# Precision Health Diagnostic and Surveillance Network uses S Gene Target Failure (SGTF) combined with sequencing technologies to identify emerging SARS-CoV-2 variants

**DOI:** 10.1101/2021.05.04.21256012

**Authors:** Rafael Guerrero-Preston, Vanessa Rivera-Amill, Karem Caraballo, Andrea Arias García, Raphael Sánchez Torres, Fernando Tadeu Zamuner, Claudio Zanettini, Matthew J. MacKay, Rachet Baits, Nike Beaubier, Gaurav Khullar, Jessica Metti, Una Pipic, Ana Purcell-Wiltz, Keilyn Vale, Gabriela Pérez, Lorena De Jesus, Yaima Miranda, Denise Ortiz, Amanda García Negrón, Liliana Viera, Alberto Ortiz, Jorge Acevedo, Josefina Romaguera, Ivonne Jiménez-Velazquez, Luigi Marchionni, José Rodríguez-Orengo, Adriana Baez, Christopher E. Mason, David Sidransky

## Abstract

Several genomic epidemiology tools have been developed to track the public and population health impact of SARS-CoV-2 community spread worldwide. A SARS-CoV-2 Variant of Concern (VOC) B.1.1.7, known as 501Y.V1, which shows increased transmissibility, has rapidly become the dominant VOC in the United States (US). Our objective was to develop an evidenced-based genomic surveillance algorithm that combines RT-PCR and sequencing technologies to identify VOCs. Deidentified data were obtained from 508,969 patients tested for COVID-19 with the TaqPath COVID-19 RT-PCR Combo Kit (ThermoFisher) in four CLIA certified clinical laboratories in Puerto Rico (n=86,639) and in three CLIA certified clinical laboratories in the US (n=422,330). TaqPath data revealed a frequency of S Gene Target Failure (SGTF) >47% for the last week of March 2021, in both Puerto Rico and US laboratories. The monthly frequency of SGTF in Puerto Rico steadily increased exponentially from 4% in November 2020 to 47% in March 2021.The weekly SGTF rate in US samples was high (>8%) from late December to early January, and then also increased exponentially through April (48%). The exponential increase in SGFT prevalence in Puerto Rico is concurrent with a sharp increase in VOCs among all SARS-CoV-2 sequences from Puerto Rico uploaded to GISAID (n=461). B.1.1.7 frequency increased from <1% in the last week of January 2021 to 51.5% of viral sequences from Puerto Rico collected in the last week of March 2021. The exponential increase in SGTF and B.1.1.7 prevalence in Puerto Rico and US requires an urgent response. According to the proposed evidence-based algorithm, approximately 50% of all positive samples should be managed as potential B.1.1.7 carriers with VOC quarantine and contact tracing protocols while their lineage is confirmed by WGS in surveillance laboratories. Patients infected with VOCs should be effectively triaged for isolation, contact tracing and follow-up treatment purposes.

## Introduction

The coronavirus disease 2019 (COVID-19), an infection caused by severe acute respiratory syndrome coronavirus 2 (SARS-CoV-2) is the first global pandemic of the 21^st^ century. Several genomic epidemiology tools have been developed to track the public and population health impact of SARS-CoV-2 community spread worldwide [1-5]. More than 137 million cases and close to 3 million deaths have been reported since the beginning of the pandemic in January 2020 (https://coronavirus.jhu.edu). A SARS-CoV-2 Variant of Concern (VOC), known as 20I/501Y.V1, VOC 202012/01, or B.1.1.7, was detected in the United Kingdom in November 2020 and has now spread to multiple countries worldwide [6-8]. Genomic epidemiology studies reveal B.1.1.7 possesses many non-synonymous substitutions of biological/immunological significance, in particular Spike mutations HVΔ69-70, N501Y and P681H, as well as ORF8 Q27stop and ORF7a [7, 9, 10]. B.1.1.7 shows increased transmissibility and has rapidly become the dominant VOC in the United States (US) (https://covid.cdc.gov) [11-14].

The HVΔ69-70 mutation is a deletion in the SARS-CoV-2 21765-21770 genome region that removes Spike amino acids 69 and 70. The HVΔ69-70 causes target failure in the TaqPath COVID-19 RT-PCR Combo Kit (ThermoFisher) assay, catalog number A47814 (TaqPath)[15]. TaqPath is designed to co-amplify sections of three SARS-CoV-2 viral genes: Nucleocapsid (*N*); Open Reading Frame 1ab (*ORF1ab*); and Spike (*S*) [16]. The Spike HVΔ69/70 deletion prevents the oligonucleotide probe from binding its target sequence, leading to what has been termed S gene dropout or S gene target failure (SGTF) [6]. SGTF is associated with significantly higher viral loads in samples tested by TaqPath[16]. S gene target late amplification (SGTL) has also been observed in a subset of samples having Cycle threshold [17] values for *S* gene >5 units higher than the maximum Ct value obtained for the other two assay targets: *N* and *ORF1ab*.

The US and countries where B.1.1.7 rapidly became the dominant SARS-CoV-2 variant require immediate and decisive action to minimize COVID-19 morbidity and mortality [14, 18]. However, the US does not have a national genomic epidemiology surveillance network for COVID-19 whole genome sequencing (WGS) program in place. Therefore, only a small fraction of all new cases is being sequenced ad-hoc. SGTF has been shown to correlate with the Δ69-70 mutation highly. Evidently, SGTF can be used as a proxy to monitor SARS-CoV-2 lineage prevalence and geo-temporal distribution and may be near-direct measure of B.1.1.7 [15, 19].

In an urgent response to the SARS-CoV-2 global pandemic, a consortium of researchers and scientists working in academia, industry, and clinical laboratories implemented a Precision Health Diagnostic and Surveillance Network (PHx) in March 2020. PHx’s original objective was to augment SARS-CoV-2 molecular testing capacity and implement a genomic surveillance network in Baltimore, New York and Puerto Rico[20]. The present work describes the development of an evidenced-based genomic surveillance algorithm that combines RT-PCR and sequencing technologies to identify VOCs.

## Materials and Methods

### Precision Health Diagnostic and Surveillance Network

The PRECEDE/PROCEED Model (PPM) [21] was selected to provide the evaluation framework for PHx conceptualization and implementation (**Supplementary Figure 1**). Weekly remote meetings began in March 2020 to perform Social, Epidemiological, Educational, Behavioral and Environmental assessments in New York, Puerto Rico and Baltimore, using the NIH I-Corps Program framework [22]. School of Medicine faculty from the University of Puerto Rico in San Juan, Johns Hopkins University in Baltimore and Weill Cornell in New York City were involved in the conceptualization and implementation of PHx. The Center for Puerto Rican Studies of Hunter College led the New York initiative. The Puerto Rico initiative was led by the Puerto Rico Public Health Trust (PRPHT) and the Baltimore initiative was led by LifeGene-Biomarks, which also coordinated the PHx consortium.

### Research samples collection

A team of investigators from the Medical Sciences Campus of the University of Puerto Rico (MSC), Johns Hopkins University School of Medicine, and LifeGene-Biomarks obtained IRB approval (IRB2770120) for a COVID-19 biomarker development study to validate TaqPath in paired, self-collected nasal swab, saliva, and urine samples. Paired samples from 280 participants were accrued from June to August 2020. Research samples were used to validate RT-PCR and isothermal (LAMP) COVID-19 clinical tests in LifeGene-Biomarks laboratory, following COVID-19 Emergency Use Authorization protocols submitted to the US Food and Drug Administration (EUA) for TaqPath. Deidentified data were used for this analysis.

### Clinical samples collection

Deidentified data were obtained from 508,969 patients tested for COVID-19 in CLIA certified clinical laboratories: 86,639 test results were from Puerto Rico and 422,330 test results were from samples collected in Connecticut, Illinois, New Jersey, and New York.

### TaqPath RT-PCR assay for detection of SARS-CoV-2

Routine clinical COVID-19 diagnostic testing was performed in Clinical Laboratory Improvement Amendments of 1988 (CLIA) regulated laboratories at University of Puerto Rico Medical Sciences Campus, Laboratorio Villa Ana, Inno Diagnostics Reference Laboratory, LifeGene-Biomarks Laboratory Yale University, Yale New Haven Hospital, and Tempus Labs, all following EUA for TaqPath COVID-19. SGTF results were defined as any SARS-CoV-2 positive sample with N or ORF1ab Ct < 30 and S gene undetermined. The data were aggregated on weekly and monthly levels for surveillance purposes.

### SARS-CoV-2 Whole Genome Sequencing

WGS data from 43,202 viral samples from Connecticut (n=3,492), Illinois (n=7,177), New Jersey (n=5,058), New York (n=27,020) and Puerto Rico (n=461), analyzed using the CoVsurver: Mutation Analysis of hCoV-19 application for mutation screening and clade classification, were downloaded on April 18, 2021, from the Global Influenza Surveillance and Response System (GISAID) site (https://www.gisaid.org).

### Biostatistics

Real-time PCR data were analyzed, interpreted and exported as .csv files using Applied Biosystems COVID-19 Interpretive Software (version 1.3). Ct values from pooled samples were removed from the data set before the analysis. Scatter plots and boxplots were prepared to visualize Ct values data. Data were summarized and correlation analyses were performed. R (version 4.0.3) was used for biostatistics analyses and data visualization. Secondary data analysis was performed on data downloaded from GISAID.

## Results

TaqPath data from close to 508,969 patients revealed a frequency of SGTF 47% for the last week of March 2021 in both Puerto Rico and US laboratories. The overall frequency of SGTL (15.1%), SGTF (9.2%), and SGTF with *N* and *ORF1ab* Cts <28 (2.5%) in Puerto Rico was high from March 2020 through March 2021. SGTF steadily increased exponentially from 4% in November 2020 to 47% in March 2021 in Puerto Rico. The average weekly SGTF rate in US samples was high (>8%) from late December to early January, and then also increased exponentially through April (48%) (**Figure 1**). During January 2021 47% of the SGTF samples were identified as B.1.1.7. when sequenced. In February, most (90%) of the sequenced samples were B.1.1.7, including 100% (147/147) from February 15-23[15].

**Figure 1.**
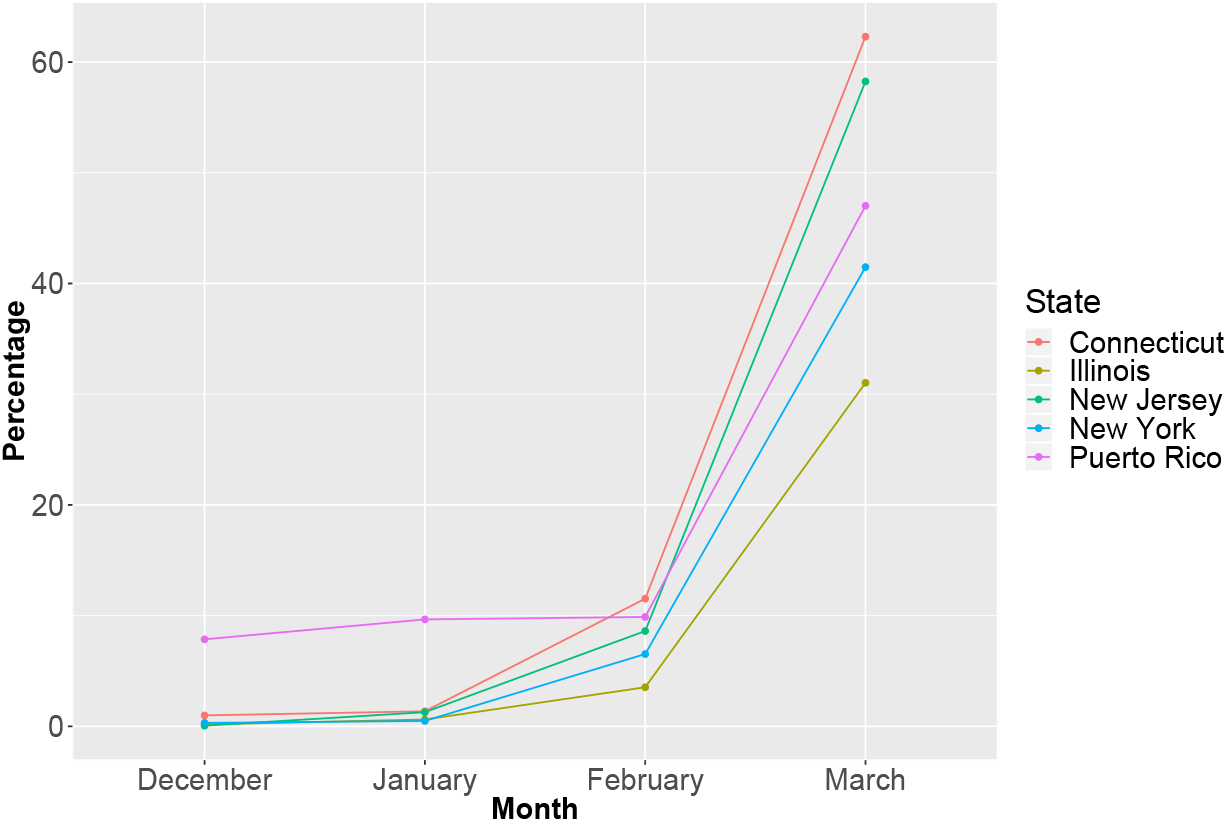
Frequency of S Gene Target Failure (SGTF) in samples from Connecticut, Illinois, New Jersey, New York, and Puerto Rico

A subset of samples (n=100) with SGTF (74%) or high viral load in *S* (<28) were evaluated with Sanger sequencing (**Supplemental Table 1**). Most samples (97%) had a Spike mutation: Δ69/70 (91%), N501Y (91%) and 82% had both Δ69/70 and N501Y. A majority of the samples (58%) with *S* <28 had the E484K mutation> E484K, a mutation located in the RBD region, is seen in SARS-CoV-2 B.1.351, P.1, P.2 and R.1 lineages[23].

The rise of SGFT in Puerto Rico is concurrent with a sharp increase in COVID-19 variants identified among viral sequences from Puerto Rico uploaded to GISAID (**Figure 2**), as of April 18, 2021 (n= 461). Viral sequences from Puerto Rico are classified under 52 different lineages and six clades Most samples were from nasopharyngeal specimens (73.9%) and are classified under seven distinct lineages: B.1.1.7 (27.7%%); B.1.588 (18.8%), B.1 (10%), B.1.2 (9.7%); B.1.1.225 (6.9%), B.1.426 (6.9%), and B.1.1 (6.4%) and four Clades: GH (49.7%); GR (21.5%); G (16.5%) and GRY (11.1%). B.1.1.7 frequency increased from <1% in the last week of January 2021, to 51.5% of viral sequences in the last week of March 2021, to become the dominant VOC In Puerto Rico.

**Figure 2.**
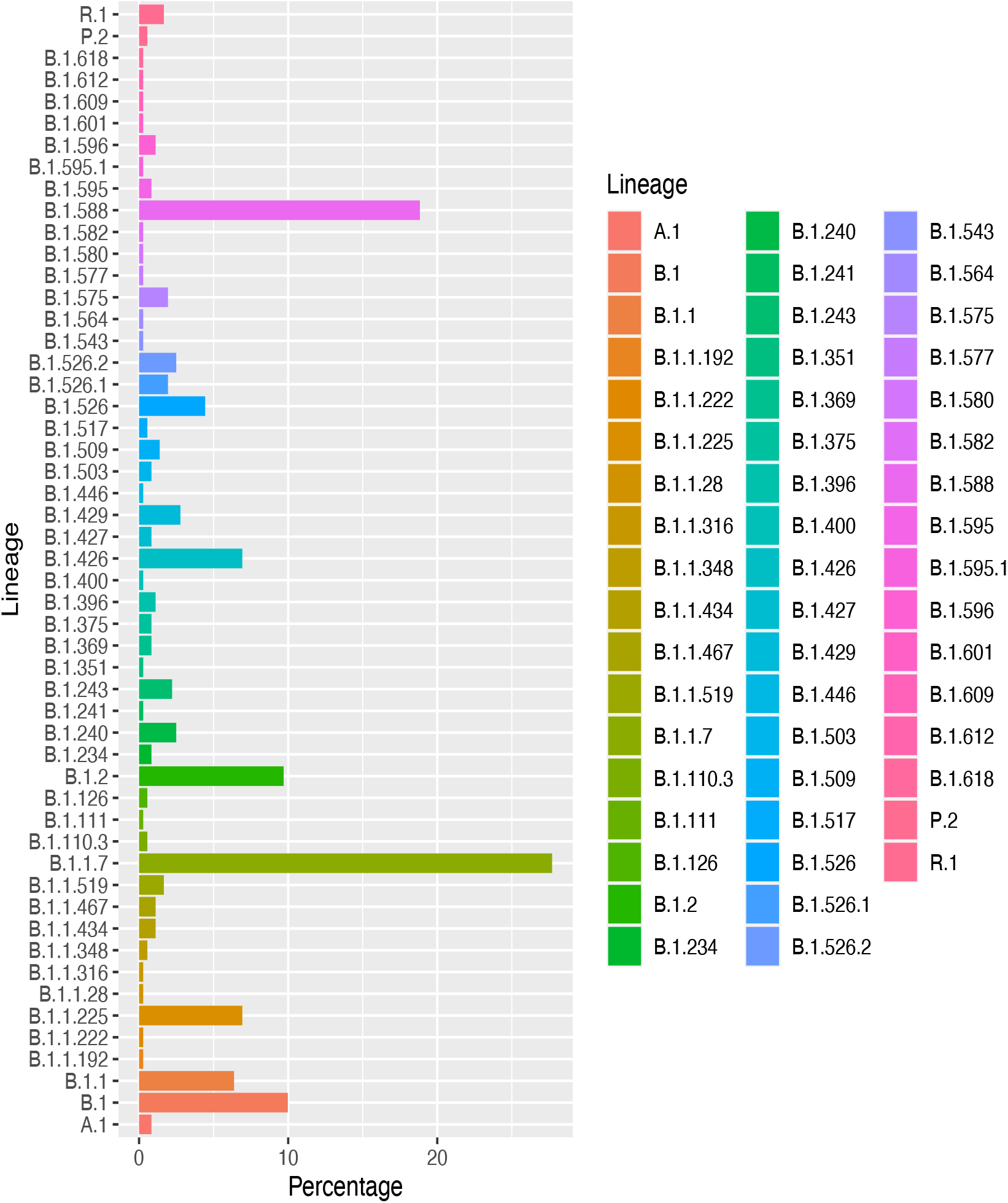
Frequency of SAS-CoV-2 variants in samples from Puerto Rico in GISAID (n=461), by Lineage.

There are 43,202 viral sequences from Connecticut, Illinois, New Jersey, New York and Puerto Rico submitted to GISAID. The median lag-time between sample collection and sample submission dates is 24 days. The lag-time mean is 51 days. The minimum lag-time is 3 days. The average daily lag-time has steadily declined from a mean of 17 days on March 1, 2021 to 14 days on March 31, 2021 (**Figure 3**).

**Figure 3.**
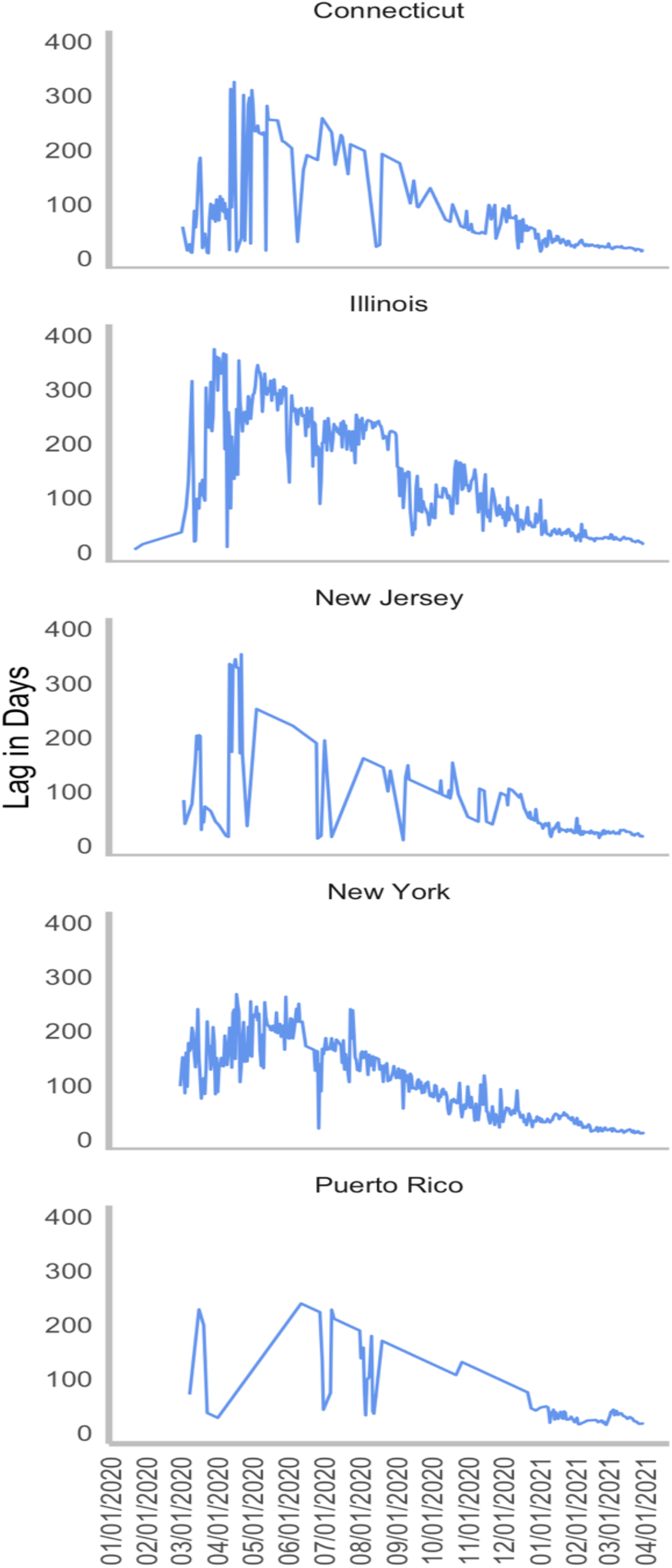
Mean daily lag time between sample collection and sequencing date from February 2020 through March 31, 2021 in Connecticut, Illinois, New Jersey, New York and Puerto Rico.

Summary statistics and Interquartile Range of *ORF1ab, N, S*, and *MS2* Cycle Threshold (Ct) [17] values for samples from Puerto Rico listed in **Table 1**, were used to develop a variant identification algorithm (**See Supplemental Materials**). The commonly used formula to calculate outliers (Q1-(1.5 x IQR) was used to identify an evidence-based Ct cutoff (Ct <28) for a proposed VOC variant identification algorithm (**Figure 4**).

**Table 1.** Summary statistics and Interquartile Range of Cycle Threshold (Ct) values for *ORF1ab, N, S, and MS2*.

| Gene | Min | Q1 | Median | Mean | Q3 | Max | NA | IQR | Outliers |
| --- | --- | --- | --- | --- | --- | --- | --- | --- | --- |
| **Puerto Rico patients (n=7,510)** |  |  |  |  |  |  |  |  |  |
| *ORF1ab* | 5.05 | 19.62 | 27.71 | 25.66 | 31.41 | 39.92 | 116 | 11.79 | 2 |
| *N* | 5.09 | 20.73 | 28.19 | 26.14 | 31.51 | 39.87 | 95 | 10.78 | 5 |
| *S* | 8.66 | 20.24 | 27.86 | 26.14 | 31.67 | 39.97 | 690 | 11.43 | 3 |
| *MS2* | 13.53 | 23.69 | 24.59 | 24.9 | 25.71 | 39.93 | 152 | 2.02 | 21 |
| **Puerto Rico patients with Ct values for *S* >= 33 and SGTF (n=1,824)** |  |  |  |  |  |  |  |  |  |
| *ORF1ab* | 5.05 | 31.97 | 33.38 | 31.91 | 34.77 | 39.17 | 100 | 2.8 | 28 |
| *N* | 5.09 | 31.94 | 33.16 | 31.75 | 34.3 | 39.87 | 63 | 2.36 | 28 |
| *S* | 33 | 33.84 | 34.83 | 35.25 | 36.2 | 39.97 | 690 | 2.36 | 30 |
| *MS2* | 13.53 | 23.82 | 24.64 | 24.78 | 25.68 | 38.83 | 15 | 1.86 | 21 |

**Figure 4.**
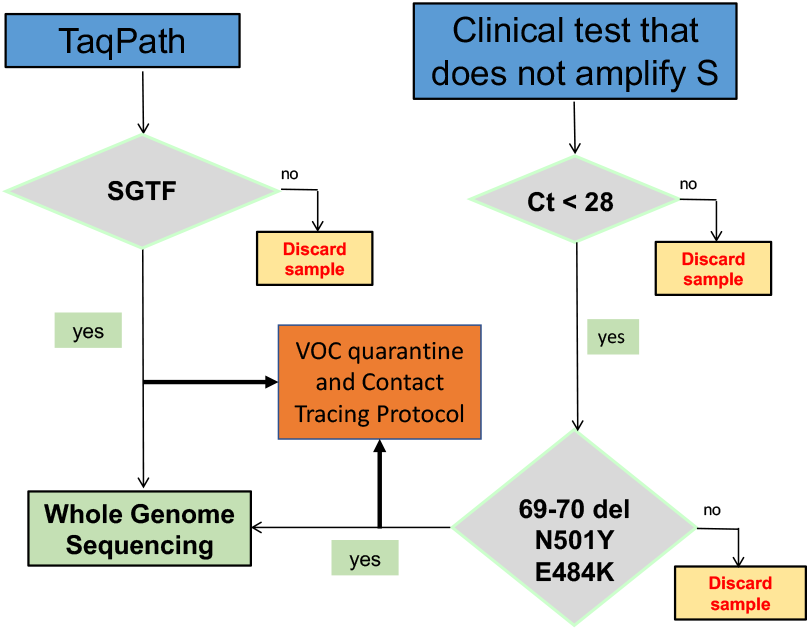
Variant of Concern Identification algorithm

Samples with SGTF should be sequenced. Samples with Ct values < 28 for *N, ORF1ab or E* in SARS-CoV-2 clinical samples without S gene data, should be evaluated for the presence of VOCs with Research use Only (ROU) kits that identify specific VOC mutations, as triage before WGS. Patients infected with VOCs should be effectively triaged for isolation, contact tracing and follow-up treatment purposes. Patients with SGTF, or positive PCR results for N501Y or E484K, should be immediately placed into quarantine and a VOC contact tracing protocol needs to be urgently set in motion. According to the proposed VOC identification algorithm, currently 50% of all positive samples in Puerto Rico and the US should be managed as potential B.1.1.7 carriers with VOC quarantine and contact tracing protocols while their lineage is confirmed by WGS in surveillance laboratories..

## Discussion

The evidence from four different US States and Puerto Rico suggests that SGTF highly correlates with B.1.1.7 prevalence. The frequency of SGTF in Puerto Rico steadily increased from 4% in November 2020 to 47% in March 2021. Similarly, SGTF in the four US States, which was high (>8%) in early January, increased to 48% in March. Concurrently, B.1.1.7 became the dominant VOC in Puerto Rico and the US in March 2021. Given the exponential rise in SGTF and B.1.1.7 prevalence, a robust VOC genomic surveillance strategy must be quickly implemented. RT-PCR can be used to identify potential VOC carriers. Patients with SGTF should be sequenced to identify viral lineage. Laboratories that do not amplify *S* can use ROU PCR or Sanger sequencing-based alternatives to identify VOC’s sentinel mutations in samples with *E, ORF1ab*, or *N* Ct values <28, as a WGS triage strategy. Patients identified as presumptive carriers of a VOCs, by SGTF or Ct values < 28, should be placed in immediate preventive quarantine until WGS data confirms their variant status.

Given the average current lag time of 15 days between clinical diagnosis and receipt of WGS results, SGTF patients should be immediately placed in quarantine for a minimum of 14 days and until they have a negative PCR test result from a self-collected sample at home. In parallel, around the clock VOC contact tracing efforts should be put in place, to prevent B.1.1.7 greater community spread. According to our evidence-based algorithm, approximately 50% of all positive patients in the four States and Puerto Rico had SGTF in March. These patients should have been placed in preventive quarantine until WGS confirmed their variant status and around the clock VOC contact tracing efforts should have been inmediately put in place.

Genomic epidemiology tools that can quickly identify and track in real-time COVID-19 VOCs improve our understanding of the transmissibility, pathogenicity, morbidity and mortality of each variant detected in geographically defined populations [24, 25]. This approach will enable the deployment of targeted, evidence-based strategies, to quickly screen for COVID-19 VOCs and identify clusters, leading to a decrease in the spread of community transmission. PHx, a critical consortium of researchers and scientists working in academia, industry, and clinical laboratories developed an evidence-based method to screen SARS-CoV-2 positive samples for COVID-19 VOCs. The PPM was a highly effective framework to guide the conceptualization and implementation of PHx. Evaluation frameworks, such as PPM, can improve the understanding of the relationship between complex variables such as community attitudes, knowledge, and screening test utilization and implementation, which determine the uptake of any screening intervention [21, 26]. Given the complexity of behavioral change processes during a global pandemic such as COVID-19, predisposing factors and barriers identified during the implementation of PHx can guide SARS-CoV-2 public policy and funding decision-making. Lessons learned from PHx can inform the urgent deployment of precision health clinical and surveillance networks. During this one year-long period the most important lesson learned is that the business model of clinical laboratories, which operate with very small profit margins, does not have much leeway for collaborative clinical or research efforts.

The main limitations of this study are the lack of established workflows, public policy guidelines and funding streams for the implementation and administration of a genomic surveillance network. The convenience samples and data used for this report was gathered ad-hoc by academic institutions, public and private organizations, as well as state and federal agencies. Data integrity, uniformity and reliability are thus compromised, and should be treated as such. Uniform sample handling and management workflows, needed to assure data reproducibility, are not in place. For example, clinical laboratories discard their samples after diagnosis, which for COVID-19 EUA approved tests, are qualitative decisions based on proprietary algorithms designed by test manufacturers. These closed PCR tests do not require Ct interpretation, nor molecular biology expertise either from the user. In addition, WGS is just entering the clinical and regulatory setting. Therefore, clinical laboratory scientists and Department of Health staff are not usually trained to sequence samples, analyze WGS data, or develop genomic surveillance programs based on RT-PCR or WGS data. The combination of these complex factors, buttressed by sample and data management asymmetry between clinical and sequencing laboratories, as well as state and federal agencies, introduce barriers to sample and data workflows, eventually impinging on results interpretation.

Our results suggest that a genomic surveillance network plays a critical role during the current stage of the COVID-19 pandemic. Patients infected with VOCs should be secured into quarantine immediately and a VOC contact tracing effort should be forcefully implemented to curtail community spread of VOCs. The evidence-based Molecular Epidemiology and Genomic Surveillance algorithm developed by PHx can quickly identify emerging VOCs as a valuable tool for identifying individual carriers of highly infectious variants, who can then be effectively triaged for isolation, contact tracing and treatment purposes.

## Data Availability

Materials availability
Data availability
Genomic data are available on GISAID (see Supplemental S3 for accession numbers).
Supplemental Information Description
Supplemental S1. Supplemental Methods, Supplemental Resutls, Supplemental Tables, Supplemental Notes.
Supplemental S2. Data for Figures 2-4.
Supplemental S3. List of SARS-CoV-2 sequences used in this study and author acknowledgements.

## Resource Availability

### Lead contact

Further information and requests should be directed to Rafael Guerrero-Preston

### Materials availability

#### Data availability

Genomic data are available on GISAID (see **Supplemental S3** for accession numbers).

### Supplemental Information Description

**Supplemental S1**. Supplemental Methods, Supplemental Resutls, Supplemental Tables, Supplemental Notes.

**Supplemental S2**. Data for Figures 2-4.

**Supplemental S3**. List of SARS-CoV-2 sequences used in this study and author acknowledgements.

## Declarations of Interests

M.J.M., R.B., N.B., G.K., J.M., C.E.M., and U.P., work for Tempus Labs. R.G.P. works for LifeGene-Biomarks. All other authors declare no competing interests.

## Acknowledgments

This research was supported by: National Institute on Minority Health and Health Disparities Small Business Innovation Research Fast Track Phase 1 / Phase 2 award (R44MD014911) and Fideicomiso de Ciencia y Tecnología de Puerto Rico Small Business Innovation Research Phase 1 Cash Match Award (R. Guerrero-Preston); MD007579: Research Centers in Minority Institutions Center for Research Resources, and Puerto Rico Science and Technology Research Trust (V. Rivera-Amill) under agreement 2020-00259; and National Cancer Institute U01CA84986 (D. Sidransky).

Thanks to Inno Diagnostics scientists: Omayra De Jesús, MT and Gerardo Hernández Buitrago, PhD; University of Puerto Rico Medical Sciences Campus Clinical Laboratory scientists: Carmen Irizarry MT and Carmen Cadilla Vázquez; Laboratorio Clínico Villa Ana scientists and staff: José Carlos Flores MBA. BS, Myrna Beltrán MPH, MT, BS; and Carla Franco MT, BS; LifeGene-Biomarks, Inc staff: Margie Cathirys Robinson, Malia Calderón and Teresa Moraima Torres; Center for Puerto Rican Studies at Hunter College Director: Edwin Meléndez, PhD; and Puerto Rico Public Health Trust scientists: Marcos López-Casillas, PhD; Puerto Rico Deparment of Health scientist: Fabiola Cruz, MS; and Center for Disease Control-Puerto Rico scientists and staff: Eddie Oneill PhD, MS, MPH and Gilberto Santiago, PhD.

## Notes

### Author Declarations

University of Puerto Rico Medical Sciences Campus (MSC) IRB approval # IRB2770120.

## References

1. Fauver, J.R., et al., Coast-to-Coast Spread of SARS-CoV-2 during the Early Epidemic in the United States. Cell, 2020. 181(5): p. 990–996 e5.

2. Zhao, Z., et al., Genetic grouping of SARS-CoV-2 coronavirus sequences using informative subtype markers for pandemic spread visualization. PLoS Comput Biol, 2020. 16(9): p. e1008269.

3. Dong, E., H. Du, and L. Gardner, An interactive web-based dashboard to track COVID-19 in real time. Lancet Infect Dis, 2020. 20(5): p. 533–534.

4. Hadfield, J., et al., Nextstrain: real-time tracking of pathogen evolution. Bioinformatics, 2018. 34(23): p. 4121–4123.

5. Meyers, L., et al., Enterovirus D68 outbreak detection through a syndromic disease epidemiology network. J Clin Virol, 2020. 124: p. 104262.

6. Borges, V., et al., Tracking SARS-CoV-2 VOC 202012/01 (lineage B.1.1.7) dissemination in Portugal: insights from nationwide RT-PCR Spike gene drop out data. Virological, 2021.

7. England, P.H., Investigation of novel SARS-COV-2 variant: Variant of Concern 202012/01: Technical briefing document on novel SARS-CoV-2 variant. [Internet]. 2020.

8. Volz, E., et al., Transmission of SARS-CoV-2 Lineage B.1.1.7 in England: Insights from linking epidemiological and genetic data. medRxiv, 2021: p. 2020.12.30.20249034.

9. Kemp, S.A., et al., Recurrent emergence and transmission of a SARS-CoV-2 spike deletion H69/V70. bioRxiv, 2021: p. 2020.12.14.422555.

10. Nemudryi, A.A., et al., SARS-CoV-2 genomic surveillance identifies naturally occurring truncations of ORF7a that limit immune suppression. medRxiv, 2021: p. 2021.02.22.21252253.

11. Washington, N.L., et al., Emergence and rapid transmission of SARS-CoV-2 B.1.1.7 in the United States. Cell, 2021.

12. Ojelade, M., et al., Travel from the United Kingdom to the United States by a Symptomatic Patient Infected with the SARS-CoV-2 B.1.1.7 Variant - Texas, January 2021. MMWR Morb Mortal Wkly Rep, 2021. 70(10): p. 348–349.

13. Washington, N.L., et al., Genomic epidemiology identifies emergence and rapid transmission of SARS-CoV-2 B.1.1.7 in the United States. medRxiv, 2021.

14. Galloway, S.E., et al., Emergence of SARS-CoV-2 B.1.1.7 Lineage - United States, December 29, 2020-January 12, 2021. MMWR Morb Mortal Wkly Rep, 2021. 70(3): p. 95–99.

15. Alpert, T., et al., Early introductions and community transmission of SARS-CoV-2 variant B.1.1.7 in the United States. medRxiv, 2021: p. 2021.02.10.21251540.

16. Kidd, M., et al., S-variant SARS-CoV-2 is associated with significantly higher viral loads in samples tested by ThermoFisher TaqPath RT-QPCR. medRxiv, 2020: p. 2020.12.24.20248834.

17. Li, H., et al., The Sequence Alignment/Map format and SAMtools. Bioinformatics, 2009. 25(16): p. 2078–9.

18. Washington, N.L., et al., Genomic epidemiology identifies emergence and rapid transmission of SARS-CoV-2 B.1.1.7 in the United States. medRxiv, 2021: p. 2021.02.06.21251159.

19. Borges, V., et al., Tracking SARS-CoV-2 lineage B.1.1.7 dissemination: insights from nationwide spike gene target failure (SGTF) and spike gene late detection (SGTL) data, Portugal, week 49 2020 to week 3 2021. Euro Surveill, 2021. 26(10).

20. Fels, J.M., et al., Genomic surveillance of SARS-CoV-2 in the Bronx enables clinical and epidemiological inference. medRxiv, 2021: p. 2021.02.08.21250641.

21. Saulle, R., et al., The PRECEDE-PROCEED model as a tool in Public Health screening: a systematic review. Clin Ter, 2020. 171(2): p. e167–e177.

22. Canaria, C.A., L. Portilla, and M. Weingarten, I-Corps at NIH: Entrepreneurial Training Program Creating Successful Small Businesses. Clin Transl Sci, 2019. 12(4): p. 324–328.

23. Huang, S.W. and S.F. Wang, SARS-CoV-2 Entry Related Viral and Host Genetic Variations: Implications on COVID-19 Severity, Immune Escape, and Infectivity. Int J Mol Sci, 2021. 22(6).

24. Volz, E., et al., Evaluating the Effects of SARS-CoV-2 Spike Mutation D614G on Transmissibility and Pathogenicity. Cell, 2021. 184(1): p. 64–75 e11.

25. da Silva Filipe, A., et al., Genomic epidemiology reveals multiple introductions of SARS-CoV-2 from mainland Europe into Scotland. Nat Microbiol, 2021. 6(1): p. 112–122.

26. Oude Munnink, B.B., et al., Rapid SARS-CoV-2 whole-genome sequencing and analysis for informed public health decision-making in the Netherlands. Nat Med, 2020. 26(9): p. 1405–1410.

