## Supplemental S1 for "Precision Health Diagnostic and Surveillance Network uses S Gene Target Failure (SGTF) combined with sequencing technologies to identify emerging SARS-CoV-2 variants"

### SUPPLEMENTAL MATERIALS

#### SUPPLEMENTAL METHODS

##### Clinical Sample collection

LifeGene provided clinical (n=3,358) and research (n=520) SARS-CoV-2 test results from samples collected from June to December 2020. Villa Ana Clinical Laboratory, a CLIA-certified community-based clinical laboratory, implemented the TaqPath assay to perform SARS-CoV-2 clinical PCR tests. Villa Ana provided 37,263 SARS-CoV-2 test results from September 1, 2020 to March 31, 2021. Inno Diagnostics Reference Laboratory and Center for Research Resources, a CLIA certified clinical and research laboratory affiliated to Ponce Medical School Foundation Inc. provided 38,794 SARS-CoV-2 test results from June 2020 to March 31, 2021. The MSC CLIA certified clinical laboratory, provided 6,704 SARS-CoV-2 test results from June 1, 2020 to March 31, 2021.

##### TaqPath RT-PCR assay for detection of SARS-CoV-2

The TaqPath™ COVID-19 Combo Kit (Applied Biosystems, Cat# A47814) was used to detect nucleic acid from SARS-CoV-2 by qualitative multiplex real-time RT-PCR, according to the manufacturer's instructions (revision F.0). Automated RNA extraction was performed using the KingFisher™ Flex Magnetic Particle Processor with 96 Deep-Well Head and the MagMAX™ Viral/Pathogen Nucleic Acid Isolation Kit (Cat# A42352) or MagMAX™ Viral/Pathogen II Nucleic Acid Isolation Kit (Cat# A48383) with a sample input volume of 200 µL. Briefly, we prepared 4 KingFisher™ Deepwell 96 Plates (Cat#  A48305) labeled: "Wash 1" (Wash buffer),

“Wash 2” (80% Ethanol), “Elution solution” and “Sample plate”. To each well of the “Sample plate", we added 5 µL of Proteinase K, 200 µL of each sample, and 200 µL of nuclease-free water to the negative control well. Binding Bead Mix previously prepared, gently mixed five times, and 275 µL added to each sample and the negative control well. Then, 5 µL of MS2 Phage control was added to each well. The MVP_2Wash_200_Flex program was used on the KingFisher™ Flex Magnetic Particle Processor with 96 Deep-Well Head (Cat# 5400630). After the run was completed, the “Elution Plate” was removed from the instrument and covered with MicroAmp™ Clear Adhesive Film (Cat# 4306311). The samples were eluted in 50 µL of Elution Solution, placed on ice for immediate use in real-time RT-PCR assay. The purified nucleic acid was reverse transcribed into cDNA and amplified using the TaqPath™ RT-PCR COVID-19 Kit. To prepare the reaction mix, we combined the following components adequate for the number of samples to be tested, in addition to  a positive control and a negative control: 6.25 µL of TaqPath™ 1-Step Multiplex Master Mix (No ROX™) (4X), 1.25 µL of COVID-19 Real-Time PCR Assay Multiplex, 7.50 µL of nuclease-free water for a total reaction mix volume of 15.0 µL. Then,  we added either 10 µL of purified sample RNA (from RNA extraction), 10 µL of Purified Negative Control, or 2 µL of Positive Control (25 copies/µL of TaqPath™ COVID-19 Control) up to 25 µL of total volume to each well of the reaction plate. We performed the RT-PCRassay using the Applied Biosystems 7500 Fast Dx Real-Time PCR Instrument, and the SDS Software v1.4.1, with the following settings: Assay: Standard Curve (Absolute Quantitation), Run mode: Standard 7500, Passive reference: None, and Sample volume: 25 µL. The data was analyzed, interpreted and exported as .csv files using Applied Biosystems COVID-19 Interpretive Software (version 1.3). R (version 4.0.3) was used for biostatistics analyses.

**SARS-CoV-2 Sanger sequencing**

A subset of SARS-CoV-2 positive clinical samples with an SGTF were selected for Sanger Sequencing in Inno Diagnostics Laboratory. Sanger sequencing primers were designed to amplify and sequence two regions of interest from the SARS-CoV-2 genome between nucleotides 21600 and 23200 of the Spike gene. Primers were designed to screen for the H69-V70 double deletion and N501Y mutations.

Samples were reverse-transcribed, and PCR amplified from the same RNA samples used for the TaqPath assay using the One Step RT-PCR kit (Qiagen). A nested PCR was performed with primers designed to identify the Δ69-70 and N501Y mutations using the FastStart PCR Master (Sigma) and confirmed using agarose gel electrophoresis and successfully amplified samples were purified. The BigDye Xterminator kit v3.1 (Thermo-Fisher) was used to purify the samples. A mix of 6.5 μL of highly deionized formamide with 3.5 μL of the purified product was used for capillary electrophoresis using a 3730xl DNA Analyzer. Electhropherogram data was aligned and quality screened to their respective region using web RECall (beta v3.05). The two reference sequences used encompass codons 30-150 and codons 417-516 of the SARS-CoV-2 Spike gene. The consensus sequences were downloaded and aligned using MegaX software.

**SARS-CoV-2 Whole Genome Sequencing in Puerto Rico**

SARS-CoV-2 Whole Genome Sequencing (WGS) was performed in a subset of samples from Puerto Rico in Ponce Research Institute. Briefly, SARS-CoV-2 WGS was completed using the Trio RNA Seq kit (Nugen Technologies) without using any human RNA depletion protocol. SARS_CoV-2 RNA purification from nasopharyngeal specimens was done using the

MagMAX™ Viral RNA Isolation Kit (Thermo Fisher Scientific) following the 200 µL purification protocol suggested for processing clinical specimens in the TaqPath™ COVID-19 Combo Kit Emergency Use Authorization. The only modification made to the protocol was the omission of the bacteriophage MS2 addition to the samples since this is a shotgun approach and competing RNA can hamper such samples' sequencing. Following RNA extraction, the samples were quantified using a Qubit 2.0 with the Qubit™ RNA HS Assay Kit (Thermo Fisher Scientific). Since the Trio RNA Seq kit can process samples in the range of 500 pg and 50 ng and none of the selected samples exceeded 2.5ng/ µL no dilution was performed. Ten µL of the RNA extraction was used to prepare the libraries, all steps for library construction were performed according to the manufacturer's protocol. The resulting libraries were quantified using the Qubit 2.0 and the Qubit™ dsDNA HS Assay Kit and diluted to a concentration of 4 nM with Tris buffer. Five µL of the diluted libraries were pooled. A final dilution to 6 pM of the libraries with a 12% PhiX spike was heat-denatured and loaded to a MiSeq 600 cycles V3 kit, and a 2 x 251 cycles run was done. After the run, the MiSeq data was analyzed using Fastqc, and the fastq files were trimmed off illumina adaptor sequences and quality filtered using Trim_galore! (https://www.bioinformatics.babraham.ac.uk/projects/trim_galore/) with the following options: --paired, -q 30, --illumina. After the quality trimming steps, each specimen fastq files were aligned to the NC_045512.2 SARS_CoV-2 reference genome using Bowtie2 [2] with –very-sensitive option and the resulting sam file was converted to bam file, sorted, and indexed using the samtools package [3].The resulting alignments were inspected using Tablet [4] for Indel verification purposes. Then, a consensus sequence was generated from the bam file using a pipe of samtools mpileup, bcftools [3] and ivar [5] packages.  The resulting consensus was analyzed using the GISAID CoVsurver: Mutation Analysis of hCoV-19 application for mutation screening and clade classification.

#### SUPPLEMENTAL RESULTS

After close to twenty remote meetings and email communications using the PPM (**Supplemental Figure 1**) a funding proposal was submitted in late April 2020 to the National Institute of Health RADx program but was not funded. The PRPHT, together with clinical Reference laboratories, began in May 2020 to coordinate SARS-CoV-2 molecular testing efforts in Puerto Rico through weekly remote meetings.

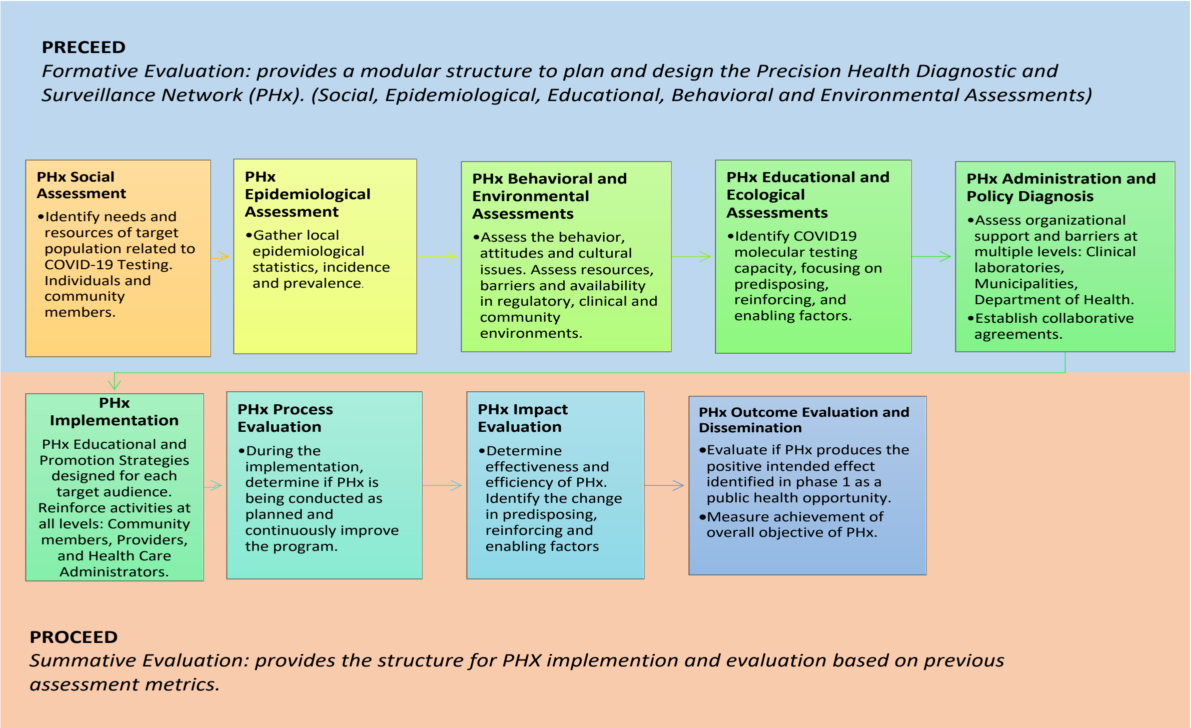


**Supplemental Figure 1.** PRECEDE/PROCEED cost–benefit evaluation framework for health promotion interventions.

In parallel, LifeGeneBiomarks began in late March 2020 to assess the possibility of opening a molecular laboratory in Puerto Rico, because all onboarding activities for new laboratories in Baltimore Bio-incubators were halted during the initial COVID19 shutdown in early March

2020. In April 2020 LifeGene and a team of researchers from the University of Puerto Rico School of Medicine obtained IRB approval to implement a Biomarker development study to compare the use of self-collected nasal, saliva and urine samples for SARS-CoV-2 diagnosis in asymptomatic participants. In June 2020 Participant accrual began in June 2020. LifeGene began testing research samples in August 2020 and clinical samples in October 2020, using the TaqPath assay.

Most (90.1%) of the positive samples (n=6,764) were identified between October 1, 2020 and March 31, 2021. Samples with an *S* Ct value >= 33, a proxy for SGTL were observed in 1,824 (24.3%) of all positive samples. SGTF was observed in 690 samples (9.2%) of all positive samples.  The frequency of SGTF steadily increased in four months from 4% in October 2020 to 47% in March 2021. SGTF rate range varied by site during these four months: MSC from 2% to 32%; Ponce from 3% to 43%; Juncos from 2% to 51%; and Vega Baja from 5% to 29%. Vega Baja did not run samples in 2021. Scatterplots and boxplots of Ct values reveal a complex

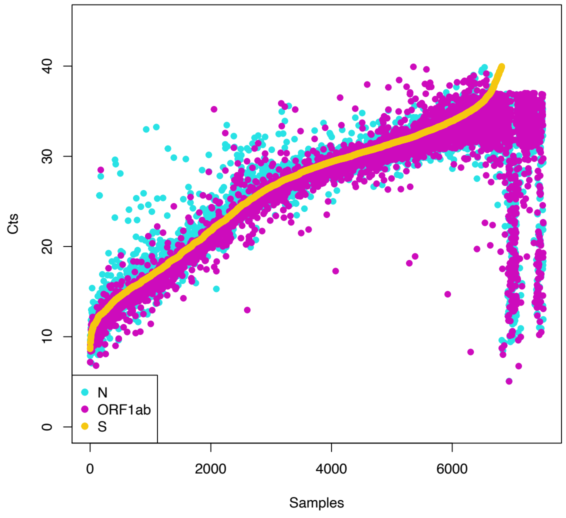


**Supplemental Figure 2a.** Cycle Threshold (Ct) values for *S, N* and *ORF1ab* in 7,510 TaqPath positive samples from Puerto Rico, diagnosed from June 2020 through March 31,2021.

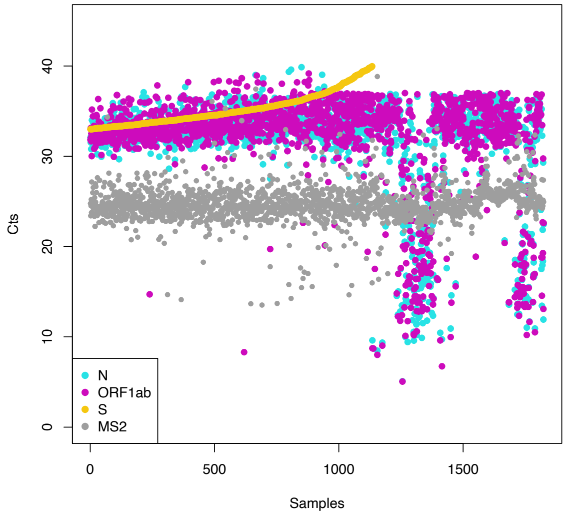


**Supplemental Figure 2b.** Cycle Threshold (Ct) values for *S, N*, *ORF1ab* and *MS2* in 1,824 TaqPath samples from Puerto Rico with S Gene Target Late Amplification (SGTL), diagnosed from June 2020 through March 31,2021.

relationship between *S, ORF1ab* and *N*, for all values of *S* (**Supplemental Figure 2a** as well as between S and *ORF1ab*, *N,* and *MS2,* when *S* >=33 (**Supplemental Figure 2b**)

A bimodal distribution of *S* Ct values is clearly apparent when comparing box plots of *ORF1ab*, *N* and *S,* for all values of *S* (**Supplemental Figure 3a**) with boxplots of *ORF1ab*, *N, MS2* and *S,* when *S* Ct >=33 (**Supplemental Figure 3b**).

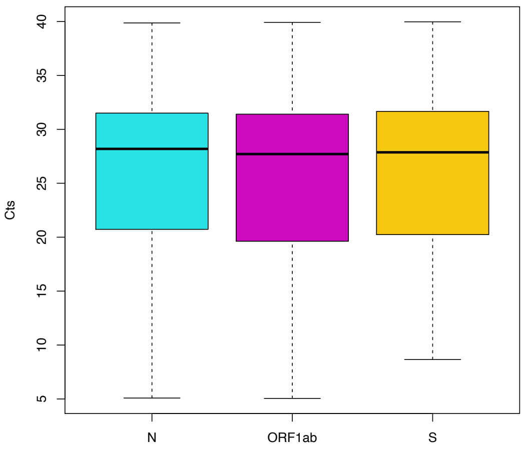


**Supplemental Figure 3a.** Box plots of Cycle Threshold (Ct) values for *S*, *N* and *ORF1ab* in 7,510 TaqPath positive samples from Puerto Rico, diagnosed from June 2020 through March 31,2021.

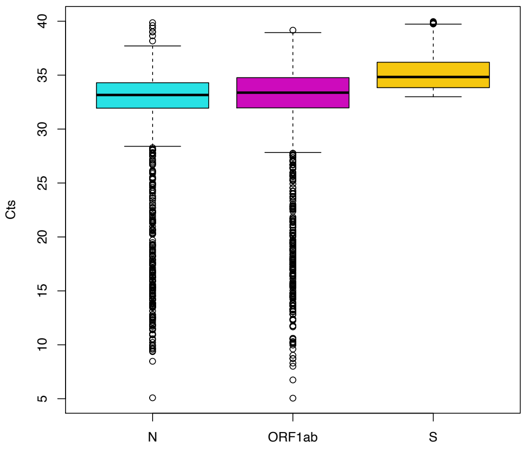


**Supplemental Figure 3b**. Box plots of Cycle Threshold (Ct) values for *S*, *N*, *ORF1ab* and *MS2* in 1,824 TaqPath samples from Puerto Rico with S Gene Target Late Amplification (SGTL), diagnosed from June 2020 through March 31,2021.

Pearson's correlation coefficient shows a highly significant correlation of *S* Ct values with *ORF1ab* (r=0.978; p<0.0001), *N* (r=0.973; p<0.0001) and a slightly inverse correlation with *MS2* (r= -0.081; p<0.0001) across the distribution of Ct values. However, sub analysis of *S* Ct values >= 33 reveals that the correlation of *S* with *ORF1ab* (r=0.169; p<0.0001) and *N* (r=0.198; p<0.0001), decreases, as the slope of the curve for *S* Ct values increases. The correlation of *S* with *MS2* Ct values was r=-0.008; p=0.79.

### SUPPLEMENTAL TABLES

**Supplemental Table 1.** Sanger sequencing results

| Study ID | Dx RT-PCR CT_Orf1ab | Dx RT-PCR CT_N | Dx RT-PCR CT_S | Sanger Sequencing (22-174) | Sanger Sequencing (391-539) |
|---|---|---|---|---|---|
| PHx-2001 | 29.4 | 29.2 | 29.9 | **N/A (Unsuccessful)** | E484K |
| PHx-2002 | 26.4 | 27.3 | 26.9 | T95I | E484K |
| PHx-2003 | 25.8 | 26.4 | 26.4 | **N/A (Unsuccessful)** | E484K |
| PHx-2004 | 25.6 | 26 | 26.2 | "T95I" | E484K |
| PHx-2005 | 25.4 | 25.8 | 26.1 | T95I | E484K |
| PHx-2006 | 25.1 | 26.6 | 26 | T95I | E484K |
| PHx-2007 | 21.9 | 22.5 | 22.4 | T95I | E484K |
| PHx-2008 | 19.8 | 20.5 | 20.8 | T95I | E484K |
| PHx-2009 | 18.3 | 19.5 | 19.9 | T95I | E484K |
| PHx-2010 | 18.3 | 19.1 | 19.1 | W152L | E484K |
| PHx-2011 | 15.7 | 15.4 | 16.2 | W152L | E484K |
| PHx-2012 | 14.6 | 15.6 | 15.4 | T95I | E484K |
| PHx-2013 | 14.7 | 14.8 | 15.3 | W152L | E484K |
| PHx-2014 | 13.7 | 14.5 | 14.6 | W152L | E484K |
| PHx-2015 | 9.4 | 10.7 | 10.6 | T95I | E484K |
| PHx-2016 | 31.2* | 31.8* | Failure* | del HV69-70 | F490L |
| PHx-2017 | 31.1* | 34.5* | Failure* | del HV69-70 | F490L |
| PHx-2018 | 26.1 | 27.2 | 26.9 | D80G, del Y144, F157S | L452R |
| PHx-2019 | 22.2 | 22.5 | 22.8 | W152C | L452R |
| PHx-2020 | 17.2 | 18.2 | 17.5 | D80G, del Y144, F157S | L452R |
| PHx-2021 | 16.4 | 17.8 | 17 | D80G, del Y144, F157S | L452R |
| PHx-2022 | 12.1 | 13.1 | 13 | D80G, del Y144, F157S | L452R |
| PHx-2023 | 31.2 | 31.6 | Failure | del H69-V70, del Y144, F157L | **N/A (Unsuccessful)** |
| PHx-2024 | 20.4 | 20.7 | Failure | del HV69-V70; del Y144 | N501Y |
| PHx-2025 | 13.8 | 13.5 | Failure | del HV69-V70; del Y144 | N501Y |
| PHx-2026 | 14.4 | 15.2 | Failure | del H69-V70, del Y144 | N501Y |
| PHx-2027 | 18 | 18.5 | Failure | del H69-V70, del Y144 | N501Y |
| PHx-2028 | 14.8 | 14.9 | Failure | del H69-V70, del Y144 | N501Y |
| PHx-2029 | 13.2 | 12.5 | Failure | del H69-V70, del Y144 | N501Y |
| PHx-2030 | 13.6 | 13.7 | Failure | del H69-V70, del Y144, S151SI | N501Y |
| PHx-2031 | 13.6 | 13.7 | Failure | del H69-V70, del Y144 | N501Y |
| PHx-2032 | 12.3 | 12.6 | Failure | del H69-V70, del Y144 | N501Y |
| PHx-2033 | 15.8 | 16.2 | Failure | del H69-V70, del Y144 | N501Y |
| PHx-2034 | 33.5 | 34.8 | Failure | **N/A (Unsuccessful)** | N501Y |
| PHx-2035 | 20.3 | 20.6 | Failure | del H69-V70, del Y144 | N501Y |
| PHx-2036 | 27.9 | 26 | Failure | **N/A (Unsuccessful)** | N501Y |
| PHx-2037 | 26.5 | 25.4 | Failure | del H69-V70, del Y144 | N501Y |
| PHx-2038 | 30.7 | 29.1 | Failure | **N/A (Unsuccessful)** | N501Y |
| PHx-2039 | 20.6 | 20.7 | Failure | del H69-V70, del Y144 | N501Y |
| PHx-2040 | 25.8 | 25.4 | Failure | del H69-V70, del Y144 | N501Y |
| PHx-2041 | 20.3 | 21.4 | Failure | del H69-V70, del Y144 | N501Y |
| PHx-2042 | 25.9 | 25.9 | Failure | del H69-V70, del Y144 | N501Y |
| PHx-2043 | 30.4 | 30.9 | Failure | **N/A (Unsuccessful)** | N501Y |
| PHx-2044 | 13.9 | 13.8 | Failure | del H69-V70, del Y144 | N501Y |
| PHx-2045 | 16.6 | 16.8 | Failure | del H69-V70, S98F, D138H, del Y144 | N501Y |
| PHx-2046 | 20.1 | 20.3 | Failure | del H69-V70, del Y144 | N501Y |
| PHx-2047 | 18.6 | 18.3 | Failure | del H69-V70, del Y144 | N501Y |
| PHx-2048 | 16.9 | 16.7 | Failure | del H69-V70, del Y144 | N501Y |
| PHx-2049 | 34.5 | 33.5 | Failure | del H69-V70, del Y144 | N501Y |
| PHx-2050 | 32.2 | 31.7 | Failure | del H69-V70, del Y144 | N501Y |
| PHx-2051 | 16.9 | 16.9 | Failure | del H69-V70, del Y144 | N501Y |
| PHx-2052 | 10.5 | 10.9 | Failure | del H69-V70, del Y144 | N501Y |
| PHx-2053 | 23.8 | 23.6 | Failure | del H69-V70, del Y144 | N501Y |
| PHx-2054 | 15.5 | 15.3 | Failure | del H69-V70, del Y144 | N501Y |
| PHx-2055 | 14.4 | 14.7 | Failure | del H69-V70, del Y144 | N501Y |
| PHx-2056 | 18.8 | 18.2 | Failure | del H69-V70, del Y144 | N501Y |
| PHx-2057 | 15.4 | 15.3 | Failure | del H69-V70, del Y144 | N501Y |
| PHx-2058 | 14.4 | 14.7 | Failure | del H69-V70, del Y144 | N501Y |
| PHx-2059 | 27.6 | 27.5 | Failure | del H69-V70, del Y144 | N501Y |
| PHx-2060 | 15 | 15.1 | Failure | del H69-V70, del Y144 | N501Y |
| PHx-2061 | 28.4 | 26.8 | Failure | del H69-V70, del Y144 | N501Y |
| PHx-2062 | 19.1 | 20.3 | Failure | del H69-V70, del Y144 | N501Y |
| PHx-2063 | 16.4 | 16.3 | Failure | del H69-V70, S98F, D138H, del Y144 | N501Y |
| PHx-2064 | 13.6 | 13.5 | Failure | del H69-V70, S98F, D138H, del Y144 | N501Y |
| PHx-2065 | 20.3 | 20.3 | Failure | del H69-V70, del Y144 | N501Y |
| PHx-2066 | 15.5 | 14.8 | Failure | del H69-V70, del Y144 | N501Y |
| PHx-2067 | 21.6 | 21.3 | Failure | del H69-V70, del Y144 | N501Y |
| PHx-2068 | 32.5 | 33.2 | Failure | **N/A (Unsuccessful)** | N501Y |
| PHx-2069 | 17.9 | 18.2 | Failure | del H69-V70, del Y144 | N501Y |
| PHx-2070 | 14.8 | 15.3 | Failure | del H69-V70, del Y144 | N501Y |
| PHx-2071 | 18.2 | 17.6 | Failure | del H69-V70, del Y144 | N501Y |
| PHx-2072 | 17.8 | 18.6 | Failure | del H69-V70, del Y144 | N501Y |
| PHx-2073 | 17.2 | 17.7 | Failure | del H69-V70, del Y144 | N501Y |
| PHx-2074 | 13.3 | 14.2 | Failure | del H69-V70, del Y144 | N501Y |
| PHx-2075 | 21.4 | 22.2 | Failure | del H69-V70, S98F, D138H, del Y144 | N501Y |
| PHx-2076 | 19.4 | 19.2 | Failure | del H69-V70, del Y144 | N501Y |
| PHx-2077 | 10.2 | 11 | Failure | del H69-V70, del Y144 | N501Y |
| PHx-2078 | 13.6 | 14.5 | Failure | del H69-V70, del Y144 | N501Y |
| PHx-2079 | 18 | 17.9 | Failure | del H69-V70, del Y144 | N501Y |
| PHx-2080 | 10.2 | 11 | Failure | del H69-V70, del Y144 | N501Y |
| PHx-2081 | 15.4 | 14.5 | Failure | del H69-V70, del Y144 | N501Y |
| PHx-2082 | 11.7 | 11.7 | Failure | del H69-V70, del Y144 | N501Y |
| PHx-2083 | 34.3 | 29.7 | Failure | **N/A (Unsuccessful)** | N501Y |
| PHx-2084 | 17.7 | 18 | Failure | del H69-V70, del Y144 | N501Y |
| PHx-2085 | 26.7 | 26.8 | Failure | del H69-V70, del Y144 | N501Y |
| PHx-2086 | 21.1 | 21.4 | Failure | del H69-V70, del Y144 | N501Y |
| PHx-2087 | 26 | 25.3 | Failure | **N/A (Unsuccessful)** | N501Y |
| PHx-2088 | 13.3 | 14.1 | Failure | del H69-V70, del Y144 | N501Y |
| PHx-2089 | 15.4 | 15.5 | Failure | del H69-V70, del Y144 | N501Y |
| PHx-2090 | 8.6 | 9.1 | 9.4 | T95I | S477N |
| PHx-2091 | 9.2 | 10.6 | 10.2 | WT | T478K |
| PHx-2092 | 34.2 | 34.4 | Failure | WT | WT |
| PHx-2093 | 26.7 | 28.1 | Failure | del HV69-70 | WT |
| PHx-2094 | 33.6 | 33.9 | Failure | WT | WT |
| PHx-2095 | 33.1 | 33 | Failure | **N/A (Unsuccessful)** | WT |
| PHx-2096 | 29.7 | 30.5 | Failure | del HV69-70 | WT |
| PHx-2097 | 29.8 | 30.4 | 39.5 | WT | WT |
| PHx-2098 | 29.7 | 30.5 | 30.4 | **N/A (Unsuccessful)** | WT |
| PHx-2099 | 15 | 16.2 | 16 | WT | WT |
| PHx-2100 | 13.5 | 13.3 | 14 | WT | WT |

| Supplemental Table 2. Changes detected through whole genome sequence analysis | | | | | | | | |
|---|---|---|---|---|---|---|---|---|
| **Virus Name** | **PR-01222021-811** | **PR-02012021-458** | **PR-02252021-211** | **PR-02252021-225** | **PR-02252021-226** | **PR-02252021-228** | **PR-02262021-383** | **PR-02262021-384** |
| **#Muts** | 30 | 27 | 16 | 15 | 14 | 14 | 33 | 32 |
| **%Muts** | 0.31% | 0.28% | 0.17% | 0.15% | 0.14% | 0.14% | 0.34% | 0.33% |
| **#UniqueMuts** | 1 | 2 | 2 | 2 | 1 | 1 | 0 | 0 |
| **%UniqueMuts** | 0.01% | 0.02% | 0.02% | 0.02% | 0.01% | 0.01% | 0.00% | 0.00% |
| **#ExistingMuts** | 29 | 25 | 14 | 13 | 13 | 13 | 33 | 32 |
| **%ExistingMuts** | 0.30% | 0.26% | 0.15% | 0.13% | 0.13% | 0.13% | 0.34% | 0.33% |
| **Clade** | GRY | GRY | GR | GR | GR | GR | GRY | GRY |
| **Lineage** | B.1.1.7 | B.1.1.7 | R.1 | R.1 | R.1 | R.1 | B.1.1.7 | B.1.1.7 |
| **GISAID Accession ID** | EPI_ISL_1201486 | EPI_ISL_1109633 | EPI_ISL_1252844 | EPI_ISL_1252845 | EPI_ISL_1235668 | EPI_ISL_1235669 | EPI_ISL_1235671 | EPI_ISL_1235672 |
| **Sample Collection Date:** | 1/22/21 | 2/1/21 | 2/25/21 | 2/25/21 | 2/25/21 | 2/25/21 | 2/26/21 | 2/26/21 |
| Reference sequence: hCoV19/Wuhan/WIV04/2019 | | | | | | | | |

Five additional samples with SGTF and Ct values for *ORF1ab* and *N* <= 30, but without Sanger sequencing results, were also selected for massively parallel sequencing [6]. Four of the five samples generated full genome sequences and were identified as SARS-CoV-2 variants of genetic lineage B.1.240 and B.1.588 (**Supplemental Table 3**)

**Supplemental Table 3.** Changes detected through whole genome sequence analysis _

| GISAID accesion # | Lineage | specimen type | symptomatic (yes/no) | Dx RT-PCR CT_MS2 | Dx RT-PCR CT_N | Dx RT-PCR CT_ORF1ab | Dx RT-PCR CT_S |
|---|---|---|---|---|---|---|---|
| EPI_ISL_1168693 | B.1.240 | saliva | no | 31.7 | 9.61 | 8.73 | SGTF |
| EPI_ISL_1168694 | B.1.240 | saliva | yes | 38.8 | 8.47 | 8.01 | SGTF |
| EPI_ISL_1168695 | B.1 | saliva | yes | 26.56 | 33.34 | 35.82 | SGTF |
| EPI_ISL_1168696 | B.1.588 | nasopharyngeal swab | yes | 26.56 | 33.34 | 35.82 | SGTF |

**SUPPLEMENTAL NOTES**

SARS-CoV-2 protein coding genes consist of seven nonstructural genes (*ORF1ab, ORF3a, ORF6, ORF7a, ORF7b, ORF8*, and *ORF10*) and four structural genes (*S, E, M*, and *N*) [7-9]. ORF1ab is the largest gene among 14 open reading frames (ORFs) encoded for 27 proteins in the SARS-CoV-2 genome. ORF1ab is located at the 5′-end of the SARS-CoV-2 genome. ORF1ab polyproteins of SARS-COV2 play an important role in viral RNA synthesis.[10] The nucleocapsid phosphoprotein, (N) coded by gene *N*, forms a capsid around the genome. The genome is further packed by an envelope coded by gene *E*, which is associated with three structural proteins: membrane protein (*M*), spike protein (*S*), and envelope protein (*E*). The N protein is highly expressed and immunogenic during infection, with the ability to enter the host cell together with the viral RNA to facilitate its replication, cooperating in the process of the virus particle assembly and release. It contains two distinct RNA-binding domains, the N-terminal domain (NTD) and the C-terminal domain (CTD), reported to bind with the viral RNA genome due to the positive amino acids and linked by a poorly structured linkage region (LKR) containing a serine/arginine-rich (SR-rich) domain (SRD).[11, 12]

S protein interacts with the host cell, and extensive structural rearrangements occur, allowing the virus to fuse with the host cell membrane. The spikes are coated with polysaccharide molecules to camouflage them, evading the host immune system's surveillance during entry[8, 9]. ORF1ab is the largest gene among 14 open reading frames (ORFs) encoded for 27 proteins in the SARS-CoV-2 genome. ORF1ab is located at the 5′-end of the SARS-CoV-2 genome. ORF1ab polyproteins of SARS-COV2 play an important role in viral RNA synthesis.

The S protein function is pivotal in the induction of neutralizing-antibody, T-cell responses, and protective immunity[13]. Mutation in the S protein (N501Y) affects the receptor-binding domain's conformation found in the B.1.1.7 variant. The same variant has 13 other B.1.1.7 lineage-defining mutations, several of which are in the S protein, including a deletion at positions 69 and 70 (del69–70) that evolved spontaneously in other SARS-CoV-2 variants and is hypothesized to increase transmissibility[14, 15]. Also, 34 deletion mutations are occurring in six sequences with 28 unique deletions in the S protein. Sequences in records MT012098 (India: Kerala State on 2020-01-27) and MT412290 (USA: WA on 2020-04-01) both have one deletion, Y145. The S protein is a type I transmembrane protein containing two subunits, S1 and S2. The first one (S1) mainly contains a receptor-binding domain (RBD) responsible for recognizing the cell surface receptor. S2 contains essential elements needed for membrane fusion. Amino acid changes at the RBD, including F490L, have been associated with augmented virus transmission in recent reports.[16]

SARS-CoV-2 clade and lineage information enables the use of sequence variation for genomic surveillance in real time. Phylodynamic thresholds, the point in time at which sufficient molecular evolutionary change has accumulated in available viral genome samples to impact transmissibility or vaccine efficacy can be used to implement evidence-based public health interventions to reduce cases and epidemic transmissibility with geo-spatial precision [17-19]. Clade and lineage information provide objective parameters to evaluate viral transmission, disease severity, and intervention effectiveness critical for guiding policy decisions that impact case burden and infection fatality ratios[20].

Clade ‘G' is the variant of the spike protein D614G which indicates significantly higher human host infectivity and better transmission efficiency to the virus. GH and GR are common offspring of clade G. According to data from GISAID, three major clades of SARS-CoV-2 are clade G (variant of the spike protein S-D614G), clade V (a variant of the ORF3a coding protein NS3-G251V), and clade S (variant ORF8-L84S).[21] Clade GH is named after the mutation in ORF3a: Q57H, and also carries mutations NSP12b: P314L and S: D614G. Clade GR carries the combination of Nucleocapsid: RG203KR, NSP3: F106F and Spike: D614G mutations. Clade O (or Others) is a general group carrying sequences which are not matching with the criteria the other clades have [22, 23].. Different fatality rates observed in different countries may be the consequence of clade's differences in virulence and immunogenicity. SARS-CoV-2 vaccines may induce clade-specific antibodies which can neutralize homologous and heterogeneous viruses with different degrees of cross-reactivity.[24-26]

Lineages are designed to capture the emerging edge of the pandemic and are at a fine-grain resolution suitable to genomic epidemiological surveillance and outbreak investigation. A lineage is as a cluster of sequences that are associated with an epidemiological event, for instance an introduction of the virus into a distinct geographic area with evidence of onward spread[23]. Lineages can be used to trace transmission chains that originate via distinct introductions from international and interstate travel, rather than widespread community transmission [27]. COVID-19 lineage evolution during the pandemic is an essential tool for the rapid detection of new SARS-CoV-2 variants needed to shape disease control and public health policies[28].

#### SUPPLEMENTAL REFERENCES


1. Washington, N.L., et al., *Genomic epidemiology identifies emergence and rapid transmission of SARS-CoV-2 B.1.1.7 in the United States.* medRxiv, 2021: p. 2021.02.06.21251159.
2. Langmead, B. and S.L. Salzberg, *Fast gapped-read alignment with Bowtie 2.* Nat Methods, 2012. **9**(4): p. 357-9.
3. Li, H., et al., *The Sequence Alignment/Map format and SAMtools.* Bioinformatics, 2009. **25**(16): p. 2078-9.
4. Milne, I., et al., *Using Tablet for visual exploration of second-generation sequencing data.* Brief Bioinform, 2013. **14**(2): p. 193-202.
5. Grubaugh, N.D., et al., *An amplicon-based sequencing framework for accurately measuring intrahost virus diversity using PrimalSeq and iVar.* Genome Biol, 2019. **20**(1): p. 8.
6. Kemp, S.A., et al., *Recurrent emergence and transmission of a SARS-CoV-2 spike deletion H69/V70.* bioRxiv, 2021: p. 2020.12.14.422555.
7. Zinzula, L., *Lost in deletion: The enigmatic ORF8 protein of SARS-CoV-2.* Biochem Biophys Res Commun, 2021. **538**: p. 116-124.
8. Telwatte, S., et al., *Novel RT-ddPCR assays for simultaneous quantification of multiple noncoding and coding regions of SARS-CoV-2 RNA.* J Virol Methods, 2021: p. 114115.
9. Telwatte, S., et al., *Novel RT-ddPCR Assays for determining the transcriptional profile of SARS-CoV-2.* bioRxiv, 2021.
10. Velazquez-Salinas, L., et al., *Positive Selection of ORF1ab, ORF3a, and ORF8 Genes Drives the Early Evolutionary Trends of SARS-CoV-2 During the 2020 COVID-19 Pandemic.* Front Microbiol, 2020. **11**: p. 550674.
11. Serrano, P., et al., *Nuclear magnetic resonance structure of the N-terminal domain of nonstructural protein 3 from the severe acute respiratory syndrome coronavirus.* J Virol, 2007. **81**(21): p. 12049-60.
12. Saikatendu, K.S., et al., *Ribonucleocapsid formation of severe acute respiratory syndrome coronavirus through molecular action of the N-terminal domain of N protein.* J Virol, 2007. **81**(8): p. 3913-21.
13. Du, L., et al., *The spike protein of SARS-CoV--a target for vaccine and therapeutic development.* Nat Rev Microbiol, 2009. **7**(3): p. 226-36.
14. Kemp, S., et al., *Recurrent emergence and transmission of a SARS-CoV-2 Spike deletion ΔH69/V70.* bioRxiv, 2020: p. 2020.12.14.422555.
15. McCarthy, K.R., et al., *Natural deletions in the SARS-CoV-2 spike glycoprotein drive antibody escape.* bioRxiv, 2020: p. 2020.11.19.389916.
16. McCarthy, K.R., et al., *Recurrent deletions in the SARS-CoV-2 spike glycoprotein drive antibody escape.* bioRxiv, 2021: p. 2020.11.19.389916.
17. Duchene, S., et al., *Temporal signal and the phylodynamic threshold of SARS-CoV-2.* Virus Evol, 2020. **6**(2): p. veaa061.
18. Oude Munnink, B.B., et al., *Author Correction: Rapid SARS-CoV-2 whole-genome sequencing and analysis for informed public health decision-making in the Netherlands.* Nat Med, 2020. **26**(11): p. 1802.
19. Lemey, P., et al., *Accommodating individual travel history and unsampled diversity in Bayesian phylogeographic inference of SARS-CoV-2.* Nat Commun, 2020. **11**(1): p. 5110.

20. Biggerstaff, M., et al., *Early Insights from Statistical and Mathematical Modeling of Key Epidemiologic Parameters of COVID-19.* Emerg Infect Dis, 2020. **26**(11): p. e1-e14.
21. Sengupta, A., S.S. Hassan, and P.P. Choudhury, *Clade GR and clade GH isolates of SARS-CoV-2 in Asia show highest amount of SNPs.* Infect Genet Evol, 2021. **89**: p. 104724.
22. Andersen, K.G., et al., *The proximal origin of SARS-CoV-2.* Nat Med, 2020. **26**(4): p. 450-452.
23. Rambaut, A., et al., *A dynamic nomenclature proposal for SARS-CoV-2 lineages to assist genomic epidemiology.* Nat Microbiol, 2020. **5**(11): p. 1403-1407.
24. Moore, J.P. and P.A. Offit, *SARS-CoV-2 Vaccines and the Growing Threat of Viral Variants.* JAMA, 2021. **325**(9): p. 821-822.
25. Soriano, V. and J.V. Fernandez-Montero, *New SARS-CoV-2 Variants Challenge Vaccines Protection.* AIDS Rev, 2021. **23**(1): p. 57-58.
26. Karim, S.S.A., *Vaccines and SARS-CoV-2 variants: the urgent need for a correlate of protection.* Lancet, 2021. **397**(10281): p. 1263-1264.
27. Deng, X., et al., *A Genomic Survey of SARS-CoV-2 Reveals Multiple Introductions into Northern California without a Predominant Lineage.* medRxiv, 2020.
28. van Oosterhout, C., et al., *COVID-19 evolution during the pandemic - Implications of new SARS-CoV-2 variants on disease control and public health policies.* Virulence, 2021. **12**(1): p. 507-508.
